# Automated Segmentation of Head and Neck Cancer from CT Images Using 3D Convolutional Neural Networks

**DOI:** 10.64898/2026.03.12.26347996

**Authors:** Piyus Prabhanjans, P Asjad Nabeel, V K Aparna, T Hannah Mary Thomas, S Balu Krishna, Hasan Shaikh, Amal Joseph Varghese, Rajendra Benny Kuchipudi, Simon Pavamani, Jeny Rajan

## Abstract

Head and neck cancer (HNC) requires accurate tumor delineation for effective radiotherapy planning. Manual segmentation of tumor regions is time-consuming and subject to considerable inter-observer variability. Although several automated approaches have been proposed, many rely on multimodal imaging such as PET/CT, which is expensive, less accessible in many clinical settings, and increases the burden on patients. In this work, we investigate a CT-only three-dimensional segmentation framework that provides a clinically practical and resource-efficient alternative. CT images of 136 head and neck cancer patients from the publicly available HN1 dataset in The Cancer Imaging Archive (TCIA) were used along with 30 additional cases from a private dataset collected at a tertiary care centre, Christian Medical College (CMC), Vellore, India. A fully automated segmentation model was developed to delineate the primary gross tumor volume (GTV) using the 3D nnU-Net framework. The models were trained using the HN1 dataset and an extended HN1+CMC dataset that included the additional private cases. Performance was evaluated using three-fold cross-validation with standard segmentation metrics including Dice Similarity Coefficient (DSC), Intersection over Union (IoU), and the 95th percentile Hausdorff Distance (HD95). The proposed CT-based model achieved a Global Dice of 0.63 and a Median Dice of 0.60 on the HN1 dataset. When the additional CMC cases were incorporated during training, the performance improved to a Global Dice of 0.65 and a Median Dice of 0.71. These results demonstrate that 3D nnU-Net can effectively segment head and neck tumors from CT images alone. The proposed CT-only approach provides a cost-effective and scalable solution that can support radiotherapy treatment planning and help reduce variability in clinical workflows.

## 1 Introduction

Head and neck cancers (HNC) present significant challenges in clinical practice due to their complex anatomical structures and the proximity of these tumors to critical organs (e.g., salivary glands, larynx, trachea, esophagus, etc.). When these tumors are treated with radiation therapy, precise outlining of the tumor boundary is essential to ensure that the maximum dose of radiation is delivered to the target while minimizing damage to surrounding healthy tissues and achieving effective tumor control [1]. Typically, tumour volumes are manually outlined (segmentation) by radiation oncologists, which is a process that is both time consuming and highly subjective. This dependence on individual expertise introduces variability and can lead to inconsistencies in treatment planning, which may result in suboptimal radiation delivery and increase the risk of tumour recurrence and side effects [2]. For instance, inadequate delineation may also contribute to downstream complications, such as patients coming back with the disease in the treated area or having side effects such as xerostomia, dysphagia, and other quality-of-life impairments that significantly affect patients undergoing treatment. These challenges highlight the need for improved methods that can enhance the accuracy and efficiency of tumor segmentation.

Automatic segmentation of head and neck tumors for treatment planning typically relies on integrating multiple modalities, such as computed tomography (CT), positron emission tomography (PET), and magnetic resonance imaging (MRI), to leverage their complementary strengths [3, 4]. CT provides detailed anatomical details, while PET provides information about the metabolic activity that can indicate tumor presence and aggressiveness. Conversely, MRI provides detailed information about soft tissue contrast, which is particularly useful to differentiate between the tumour and its surrounding healthy tissue.

Deep learning methods, particularly convolutional neural networks (CNNs) and other advanced architectures, have emerged as powerful tools for the automated segmentation of head and neck cancer from acquired images [5]. These techniques significantly outperform conventional methods by enabling rapid, high-precision segmentation that accounts for patient-specific tumor characteristics. By learning from large, annotated imaging datasets, deep learning models can detect subtle variations in tissue contrast and tumor boundaries, offering greater accuracy and consistency than manual segmentation. Recent advancements, such as hybrid architectures and U-Net-based models, have shown exceptional efficacy in capturing the complex morphology of HNC tumors on CT and PET/CT scans [6], facilitating accurate tumor delineation even in challenging cases.

However, in low and middle-income countries (LMICs) and other resource-limited settings, access to advanced imaging technologies such as PET and MRI for head and neck cancers may be limited, often restricting clinicians to using only CT scans for treatment planning [7]. To address these challenges, our aim was to develop a fully automated segmentation model utilizing a self-configurable nnU-Net deep learning framework, specifically designed for CT images. nnU-Net was chosen due to its ability to automatically adapt to diverse datasets without requiring extensive manual intervention [8]. The framework systematically analyzes the provided training cases, creating tailored configurations that optimize the segmentation process based on the unique characteristics of the data set. Focusing on the widely available CT modality, our approach aims to enhance the accuracy and efficiency of tumor delineation for head and neck cancers, particularly in low-resource settings.

The key contributions of this study are as follows.

- Development of a fully automated CT-based segmentation model to segment the HNC primary gross tumor volumes using the 3D nnU-Net framework.
- Evaluation of the model using three-fold cross-validation on the public HN1 dataset and an extended dataset with 30 additional private cases from Christian Medical College (CMC), Vellore, demonstrating performance improvements.
- A cost-effective and scalable CT-only approach that can be applied in resource-limited clinical workflows.

## 2 Related Work

Recent advances in deep learning-based segmentation of head and neck tumors using multimodal imaging have shown great promise. However, segmenting these tumors on CT images remains a significant challenge. Oreiller et al. [9] introduced the first public Head and Neck Tumor segmentation challenge (popularly known as HECKTOR), specifically delineating oropharyngeal primary tumors on FDG-PET/CT images. The best performing model used U-Net architecture with squeeze-and-excitation normalization blocks, achieving an average Dice Similarity Coefficient (DSC) of 0.76. Meanwhile, Berthon et al. [10] proposed a decision tree-based supervised learning algorithm for PET imaging, which attained a mean DSC of 0.82. Moe et al. [11] explored 2D-CNN models for segmenting both gross tumor and nodal volumes, employing four different loss functions including cross-entropy, Dice, and *f*_*β*_ loss with *β* ∈ {2, 4}. The highest DSC achieved in their study was 0.71.

Guo et al. [12] proposed a DenseNet framework to enhance information propagation and leverage features from multi-modal input images, and compared it with the 3D U-Net segmentation task. Additionally, they analyzed the performance differences between single-modal input (PET or CT) and multi-modal input (PET/CT) for each frame. Their proposed multi-modal DenseNet outperformed the reference network, achieving a Dice score of 0.73 compared to 0.71. Furthermore, the DenseNet framework required fewer trainable parameters than the 3D U-Net, reducing model complexity while maintaining superior performance.

Ren et al. [13] proposed a residual 3D U-Net architecture for delineating primary tumors on CT, PET, and MRI scans, as well as their combinations (PET-MRI, MRI-PET-CT, PET-CT, and CT-MRI). To address the class imbalance problem, they employed a combination of Dice and focal losses as their loss function. Their PET-CT-based model achieved the highest DSC of 0.77. Similarly, Naser et al. [14] reported a DSC of 0.77 while using a 3D Residual U-Net (ResUnet) framework on PET-CT images. Moe et al. [11] evaluated CNNs for auto-delineating GTV in head and neck cancer (HNC) using multimodal PET/CT imaging. Their PET/CT-based models achieved a mean Dice score of 0.71, outperforming CT-only models (0.56) and earlier studies (0.31). The study emphasized the importance of preprocessing CT images with a soft-tissue window (− 30 to 170 Hounsfield Units) to enhance performance. They highlighted the need for multi-center validation and improved artifact management for broader applicability in radiotherapy planning.

Gay et al. [15] performed automatic segmentation of primary and nodal tumor volumes on contrast-enhanced CT and synthetic MRI images using the nnU-Net architecture and five different techniques. They reported that specialized architectures, such as attention-gated U-Net, did not provide significant improvements over nnU-Net. The best performance was achieved using contrast-enhanced CT images, with the highest median DSC of 0.70. However, consistency remained a challenge, particularly in nodal tumor identification.

Liedes et al. [16] explored 2D U-Net models for segmenting head and neck squamous cell carcinoma (HNSCC) using PET and MRI. The best results were achieved with the combined PET-MRI model, which yielded a mean DSC of 0.87. Zhao et al. [17] implemented a 3D U-Net model using both CT and PET. Their architecture incorporated five encoding-decoding levels with deep supervision, a channel dropout strategy to handle missing modalities, and a combination of conventional and dilated convolutions to capture both fine details and global context. The combined PET-CT model achieved the highest DSC of 0.80.

Reinders et al. [18] investigated the use of nnU-Net for automatic lymph node segmentation in HNSCC using MRI. They developed two models: one for delineating lymph node levels, verified by a radiation oncologist, and another for segmenting individual nodes. Their approach achieved a median DSC of 0.72 for elective lymph nodes, outperforming manual segmentation in recall (*p* = 0.002). Despite these promising results, challenges remained, including interobserver variability, suboptimal image quality in certain regions, and the need for retraining to accommodate different scanner protocols.

In general, while PET /CT-based models have demonstrated higher segmentation performance, they may not be available in low-resource settings. In addition, there is a research gap when it comes to using CT alone, which has the potential to overcome these challenges effectively. Another critical observation was the limited use of 3D configurations in existing models, despite their potential to improve accuracy and overall performance. Building on these insights, our work focuses on developing a fully automated end-to-end 3D segmentation model that uses CT imaging only to enhance segmentation accuracy and reliability in the planning of radiotherapy for head and neck cancer.

## 3 Methods

### 3.1 Dataset

For this study, we used two datasets: a public dataset, *HEAD-NECK-RADIOMICS-HN1*, from The Cancer Imaging Archive (TCIA) [19], comprising 137 patients, and a private institutional dataset named *HNC*, consisting of 30 patients. The private data was collected at Christian Medical College (CMC) Vellore and was ethically approved by the institutional review board. All CT volumes were de-identified prior to use and all patients had histologically confirmed head and neck squamous cell carcinoma and were treated with radiation therapy and chemotherapy.

Due to technical issues, one patient from the HN1 dataset was excluded, resulting in a total of 136 usable scans. To ensure a robust evaluation and minimize the risk of overfitting, a three-fold cross-validation strategy was adopted. In each fold, the model was trained and validated on distinct subsets and tested on entirely unseen patients. This approach provided a reliable estimate of model performance and improved generalizability across the HN1 dataset, the CMC dataset, and their combined cohort, reflecting the model’s adaptability to variations in data from different clinical sources. The patient selection and splitting process is summarized in Figure 1. Representative examples from both datasets are shown in Figure 2.

**Figure 1:**
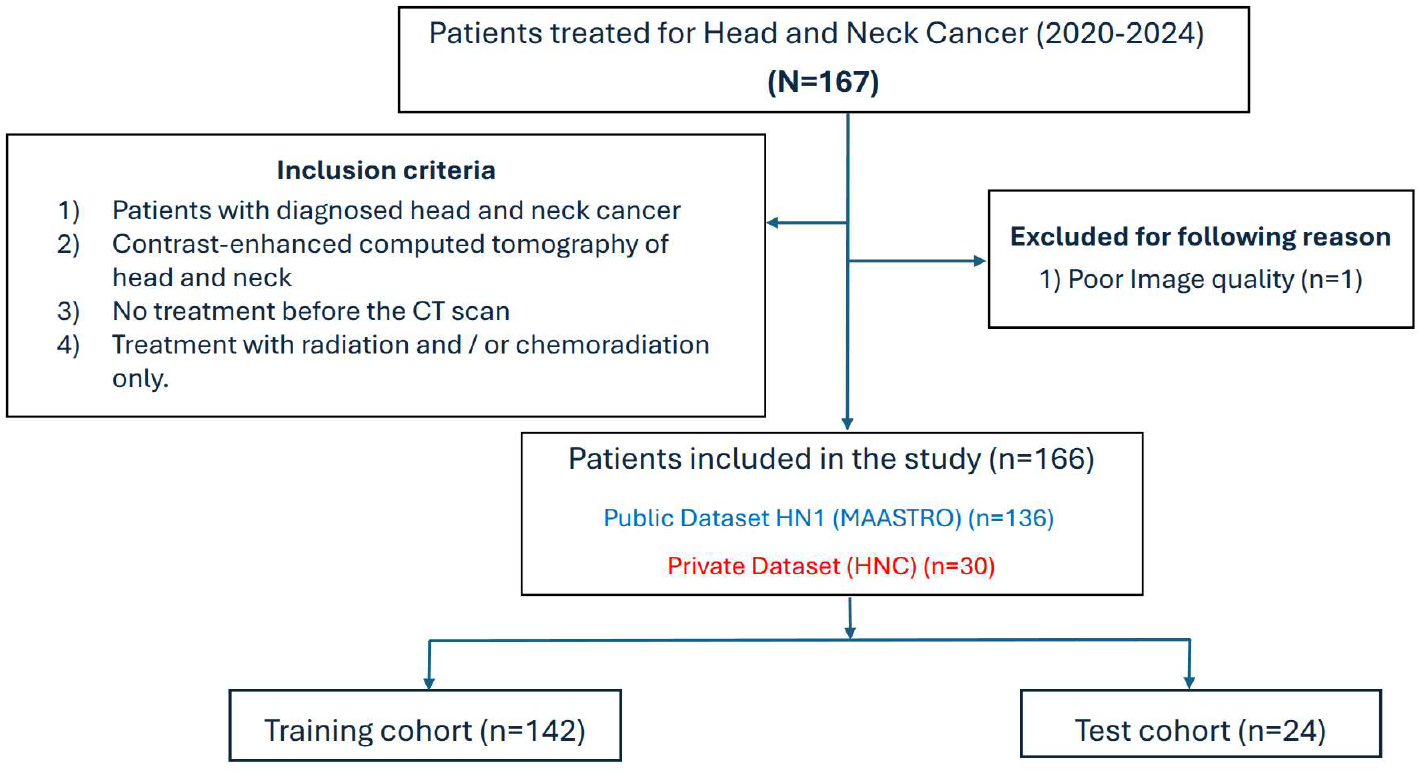
Patient selection and dataset splitting flowchart

**Figure 2:**
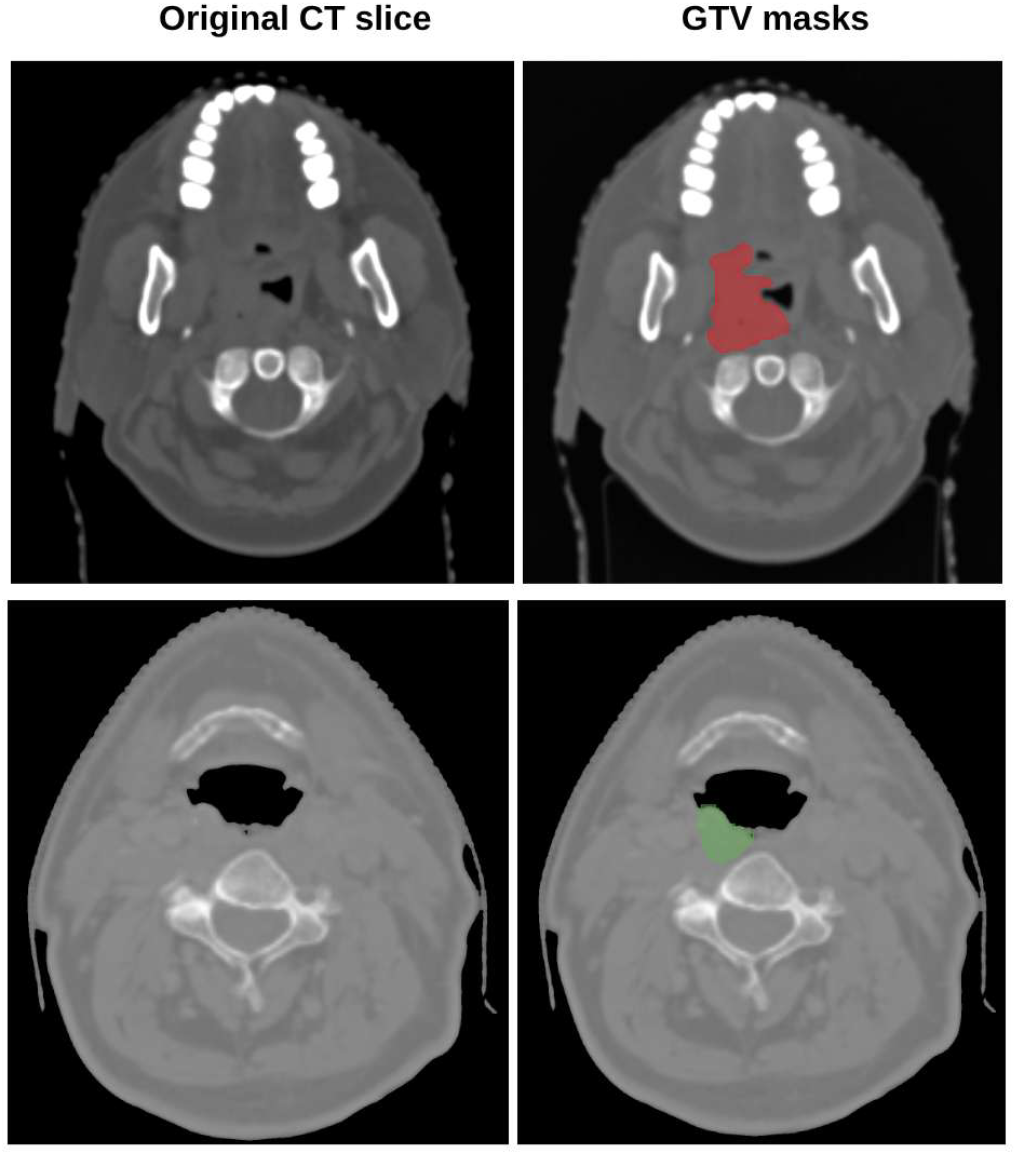
Example CT slices with gross tumor volume (GTV) masks from the private HNC dataset (top row) and the public HN1 dataset (bottom row). Left column: original CT slice; Right column: segmented GTV mask

All CT scans were contrast enhanced and had an in-plane resolution of 512 × 512 pixels with slice thickness of 2-3mm. The gross tumour volumes (GTVs) regions were manually annotated by experienced radiation oncologists at the correpsonding centers and were saved in DICOM RT Struct format. All volumes were reviewed by an independent radiation oncologist from CMC to ensure only the primary tumour was included.

### 3.2 Preprocessing

The CT images and corresponding masks were preprocessed to ensure consistent input quality. The CT volumes were resampled to an isotropic voxel spacing of 1.0 mm × 1.0 mm × 1.0 mm along the *x, y*, and *z* axes. The tumour outlines were converted into binary masks for the segmentation pipeline. Each CT was then cropped to 256 × 256 pixels, centered on the head and neck region, to remove irrelevant structures such as the imaging couch and surrounding air.

To enhance soft-tissue contrast, a fixed window level (WL) of 40 and a window width (WW) of 400 were applied, in line with radiological standards for head and neck imaging shown in Figure 3. This ensured that tumor boundaries were clearly distinguished from surrounding muscle and bone. Intensity normalization was performed using a z-score approach, where voxel values were standardized based on the mean and standard deviation of non-zero regions:

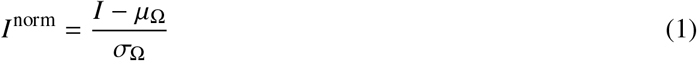

where Ω denotes the set of non-zero voxels per scan. This reduced inter-patient variability caused by differences in acquisition protocols and stabilized network optimization.

**Figure 3:**
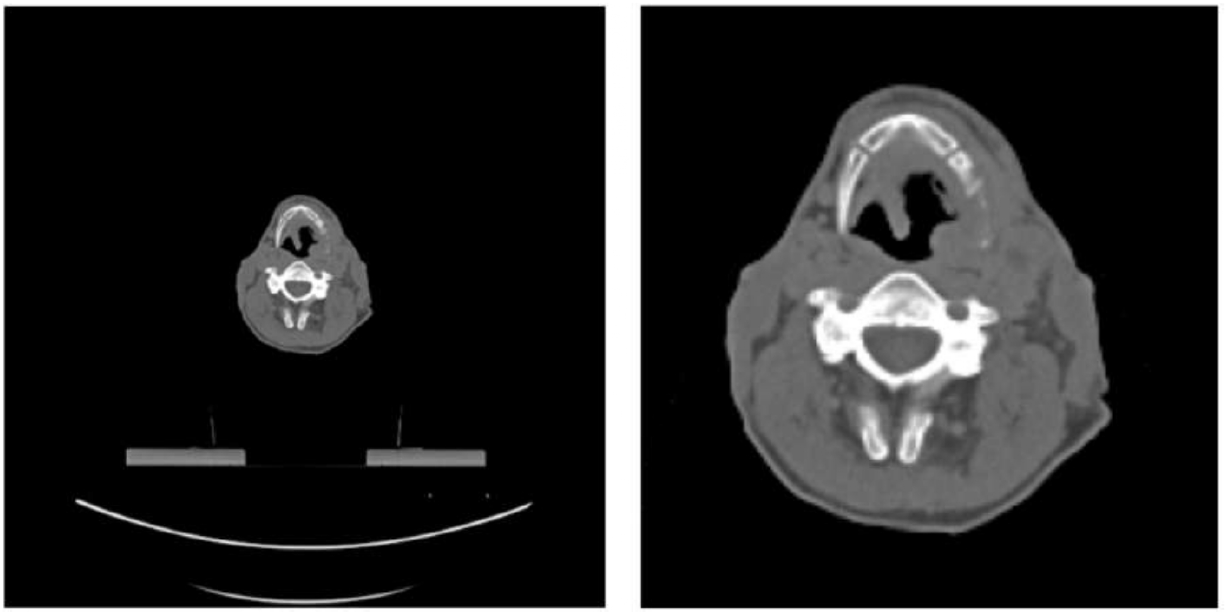
Comparison of CT images before (left) and after preprocessing (right), showing the enhanced region of interest for segmentation

### 3.3 Model Training

The proposed segmentation model uses a 3D nnU-Net v2 framework which autonomously derives dataset-specific hyperparameters collectively referred to as the *data fingerprint*. This includes characteristics such as image dimensions, voxel spacing, and modality-specific features. These inferred parameters, in conjunction with fixed components including the network architecture and loss function, form a comprehensive *pipeline fingerprint* that governs the training process. Figure 4 provides an overview of the segmentation pipeline.

**Figure 4:**
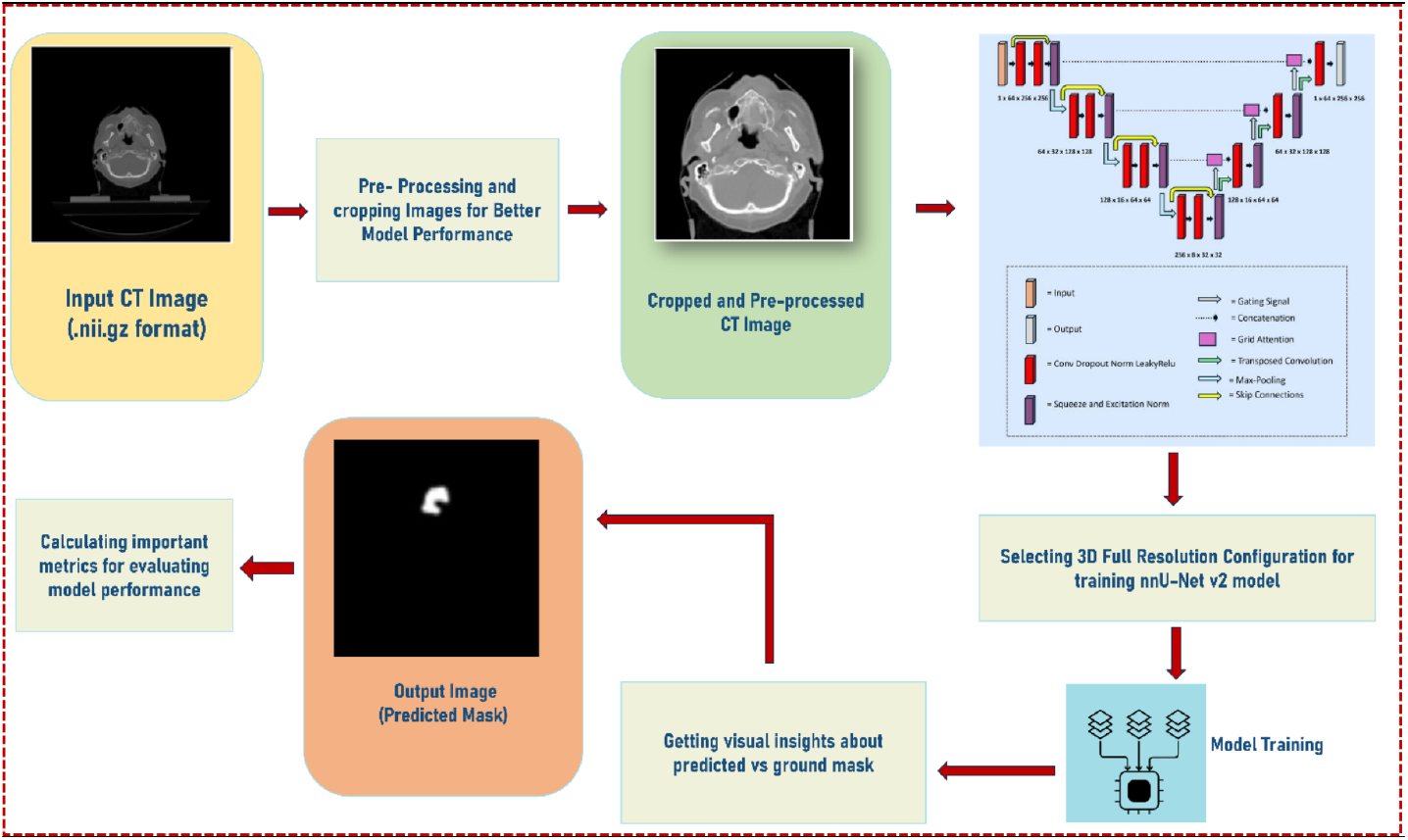
Proposed segmentation pipeline

The 3D variant of nnU-Net was selected as it offers distinct advantages over its 2D counterpart, including richer contextual information, superior spatial representation, and enhanced segmentation accuracy for volumetric medical images which enhances in segmentation performance, particularly when applied to anatomically complex and large-scale data.

The underlying architecture of nnU-Net [20] adopts an encoder–decoder structure with skip connections, which facilitates the preservation of spatial context while enabling efficient feature propagation across layers. The network leverages multi-resolution feature extraction to capture fine-to coarse-level details, deep supervision to strengthen learning in intermediate layers, and automated hyperparameter tuning to maximize segmentation effectiveness.

In this study, we adopted the FULL Resolution configuration (`3D_FullRes`) of nnU-Net v2 to retain high-fidelity anatomical details from the CT images. To evaluate the impact of architectural scaling, we conducted additional experiments with residual encoder configurations, specifically the MEDIUM (M) and LARGE (L) variants. Model optimization was performed using Stochastic Gradient Descent (SGD) with Nesterov momentum, ensuring stable convergence and efficient training dynamics. The final configuration was selected based on a comparative analysis of segmentation performance metrics. The total loss *L*_total_ is computed as the sum of Dice loss *L*_dice_ and Cross-Entropy loss *L*_CE_, defined as:

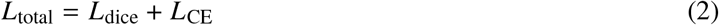

The Dice loss *L*_dice_ focused on the region-level overlap between the predicted segmentation mask *ŷ* and the ground truth mask *y*, and is given by:

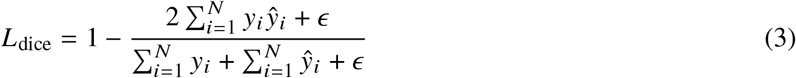

where, *y*_*i*_ and *ŷ*_*i*_ represent the ground truth and predicted values for voxel *i*, respectively. *N* denotes the total number of voxels, and *ϵ* is a small constant added for numerical stability to prevent division by zero. Dice loss directly optimizes the overlap between predicted and actual segmentation masks, making it particularly effective for segmenting smaller or less prominent tumour regions and for handling class imbalance.

The Cross-Entropy loss *L*_CE_ is defined as:

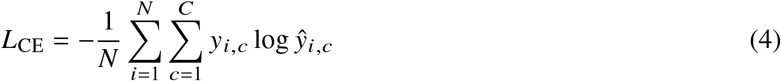

Here, *C* is the number of classes, and *y*_*i,c*_ and *ŷ*_*i,c*_ represent the true and predicted probabilities for voxel *i* belonging to class *c*. This loss function penalized misclassifications at the pixel level, encouraging sharper and more accurate boundary predictions.

Together, these losses complemented each other: Dice loss enhanced the region-level segmentation accuracy, while Cross-Entropy loss sharpened pixel-wise distinctions, especially around tumour boundaries. This combined loss function was applied at the output of every decoder block within the nnU-Net framework, leveraging deep supervision to improve gradient flow and ensure accurate segmentation across all layers.

The hyperparameters for the 3D segmentation model were carefully selected through a series of controlled experiments, balancing empirical evidence from prior literature and systematic tuning on our dataset. We initially experimented with different numbers of training epochs (500, 1000, and 2000) to identify a value that ensures stable convergence without unnecessary overfitting. Based on these trials, 2000 epochs were chosen as an optimal balance between performance and computational cost. A relatively high initial learning rate of 1 × 10^−2^ was adopted to accelerate convergence during early training, complemented by a weight decay of 3 × 10^−5^ to reduce overfitting and ensure generalization. Each training epoch consisted of 250 iterations, while 50 validation iterations were performed to provide a trade-off between efficient training and reliable monitoring of model performance. Key hyperparameters are listed in Table 1; the network stages are summarized in Table 2.

**Table 1:** Hyperparameters used for training the Proposed 3D segmentation model.

| Hyperparameter | Value |
| --- | --- |
| Initial learning rate | $1 \times 10^{-2}$ |
| Weight decay | $3 \times 10^{-5}$ |
| Oversample foreground percent | 0.33 |
| Probabilistic oversampling | False |
| Iterations per epoch (training) | 250 |
| Iterations per epoch (validation) | 50 |
| Number of epochs | 2000 |
| Deep supervision | Enabled |

**Table 2:** 3D nnU-Net based network architecture for HNC Segmentation.

| Stage | Operations | Kernel | Stride | Ch./Out. |
| --- | --- | --- | --- | --- |
| Enc-1 | Conv3D+IN+LReLU×2 | 3×3×3 | 1×1×1 | 32 |
| Enc 2–4 | Down+Conv×2 (rep.) | 3×3×3 | 2×2×2 | 64/128/256 |
| Enc-5 | Down+Conv×2 | 3×3×3 | 2×2×1 | 320 |
| Enc-6 | Down+Conv×2 | 3×3×3 | 2×2×1 | 320 |
| Bottleneck | Conv×2 | 3×3×3 | 1×1×1 | 320 |
| Dec-6 | Up+Skip+Conv×2 | 3×3×3 | 2×2×1 | 320 |
| Dec-5 | Up+Skip+Conv×2 | 3×3×3 | 2×2×1 | 320 |
| Dec 4–2 | Up+Skip+Conv×2 (rep.) | 3×3×3 | 2×2×2 | 256/128/64 |
| Dec-1 | Up+Skip+Conv×2 | 3×3×3 | — | 32 |
| Output | Conv3D 1×1×1 + Softmax | 1×1×1 | — | 224×192×56 |

Foreground oversampling was incorporated because the tumor regions occupy a relatively small fraction of the total CT volume. Several experiments were conducted with different oversampling foreground ratios ranging from 0.30 to 0.40, and a value of 0.33 provided the best trade-off between learning efficiency and segmentation accuracy. Deep supervision was enabled to propagate meaningful gradients to intermediate layers of the network, encouraging multi-scale feature learning and improving segmentation accuracy on fine tumor boundaries. These hyperparameters represented the outcome of a structured exploration process designed to mitigate common challenges in 3D medical image segmentation, including class imbalance, convergence instability, and overfitting, ultimately leading to a robust and reproducible training setup.

In this study, the model was applied to head and neck CT scans with a median axial resolution of 510 × 510 and a depth of 134 slices. Based on dataset statistics and memory constraints, nnU-Net selected an input patch size of 224×192×56 voxels to ensure efficient training while maintaining sufficient spatial context. Rather than processing entire volumes, the network performed inference using overlapping patches and a sliding window strategy. Patch-wise predictions were aggregated through Gaussian-weighted soft-voting, resulting in full-volume segmentations of shape *C* × 510 × 510 × 134, where *C* denotes the number of segmentation classes.

The architecture followed a six-stage encoder–decoder design. Each encoder stage consisted of two convolutional blocks comprising a 3D convolution layer (kernel size 3 × 3 × 3), instance normalization, and in-place LeakyReLU activation. The number of feature channels increased with depth: 32, 64, 128, 256, and 320. Spatial resolution was reduced using strided convolutions symmetric strides (2 × 2 × 2) in the early stages and asymmetric strides (2 × 2 × 1) in the deeper layers to preserve z-axis resolution. The decoder mirrored the encoder in depth and structure, using transposed convolutions for upsampling and skip connections to fuse encoder features at corresponding resolution levels. Deep supervision was applied at intermediate decoder outputs to aid convergence. A final 1 × 1 × 1 convolution followed by softmax activation generated per-patch prediction. These patches were augmented back to the original CT dimensions to get the predicted results.

### 3.4 Evaluation

The evaluation metrics included Global Dice, Median Dice, Precision, Sensitivity, Specificity, Intersection over Union (IoU), and 95th Percentile Hausdorff Distance (HD95). We report fold-wise values and the mean across folds. All experiments were conducted on an NVIDIA workstation equipped with an NVIDIA L40 Tensor Core GPU (48 GB memory), using CUDA Toolkit 12.6.0, Python 3.9+, and PyTorch.

## 4 Results

### 4.1 Quantitative Results

Table 3 summarizes the performance across three folds of the combined HN1+CMC dataset, along with the HN1-only baseline for reference.

**Table 3:**
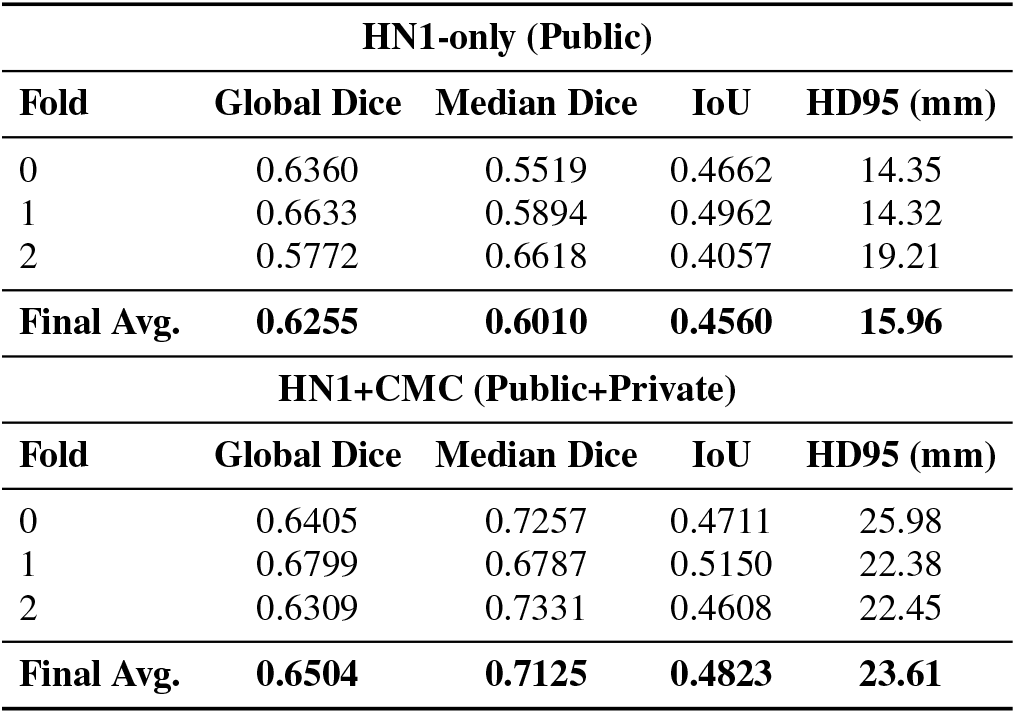
Segmentation performance across three folds on HN1-only (Public) and HN1+CMC (Public+Private) datasets.

| HN1-only (Public) |  |  |  |  |
| --- | --- | --- | --- | --- |
| Fold | Global Dice | Median Dice | IoU | HD95 (mm) |
| 0 | 0.6360 | 0.5519 | 0.4662 | 14.35 |
| 1 | 0.6633 | 0.5894 | 0.4962 | 14.32 |
| 2 | 0.5772 | 0.6618 | 0.4057 | 19.21 |
| **Final Avg.** | **0.6255** | **0.6010** | **0.4560** | **15.96** |
| HN1+CMC (Public+Private) |  |  |  |  |
| Fold | Global Dice | Median Dice | IoU | HD95 (mm) |
| 0 | 0.6405 | 0.7257 | 0.4711 | 25.98 |
| 1 | 0.6799 | 0.6787 | 0.5150 | 22.38 |
| 2 | 0.6309 | 0.7331 | 0.4608 | 22.45 |
| **Final Avg.** | **0.6504** | **0.7125** | **0.4823** | **23.61** |

Compared to the HN1-only baseline, the inclusion of CMC data (averaged across all three folds) resulted in a relative improvement of +3.99% in Global Dice (0.6255 → 0.6504), +18.56% in Median Dice (0.6010 → 0.7125), and +5.77% in IoU (0.4560 → 0.4823). However, the HD95 metric increased from 15.96 mm to 23.61 mm, indicating reduced boundary precision. Fold-wise analysis further revealed that Fold 1 benefited the most from the additional data, while Folds 0 and 2 exhibited only marginal changes, underscoring the impact of case distribution and data heterogeneity on model generalization.

### 4.2 Results of HN1 Public Dataset

Experiments conducted exclusively on the HN1 public dataset established the baseline performance of the proposed segmentation framework. Across three folds, the model achieved average scores of 0.6255 for Global Dice, 0.6010 for Median Dice, and 0.4560 for IoU, reflecting moderate overlap between predicted tumor masks and ground truth annotations. Precision remained consistently high (0.8603–0.8724), suggesting that the network effectively minimized false positives. However, sensitivity values were lower (0.4472–0.5351), indicating that small or diffuse tumor regions were often under-segmented. This trade-off between high precision and limited sensitivity highlights the difficulty of capturing the full extent of irregularly shaped tumors in head and neck CT scans. Specificity, as expected, remained close to unity across folds, owing to the dominance of non-tumor background voxels.

Boundary accuracy analysis further supported these findings. The HD95 values ranged between 14.32 mm and 19.21 mm, with an average of 15.96 mm, indicating stable alignment between predicted and reference contours in most cases. Fold-wise variation was evident, with Fold 2 showing the lowest Global Dice (0.5772) but relatively higher Median Dice (0.6618), suggesting that case distribution and tumor characteristics influenced fold performance differently. Overall, the HN1-only experiments demonstrated that the model is capable of generating robust and precise segmentations on a large public dataset, but challenges remain in maximizing sensitivity and fully capturing tumor boundaries.

### 4.3 Overall Comparison Across Metrics

Figure 5 illustrates segmentation performance across the three folds, using two rows of boxplots: the first row presents region-overlap measures (Global Dice, Median Dice, and IoU), while the second row summarizes classification- and distance-based measures (Precision, Sensitivity, and HD95). Together with Table 3, these plots provide a comprehensive overview of model behavior, highlighting fold-to-fold variability and enabling intuitive interpretation of volumetric overlap, voxel-wise classification accuracy, and boundary precision.

**Figure 5:**
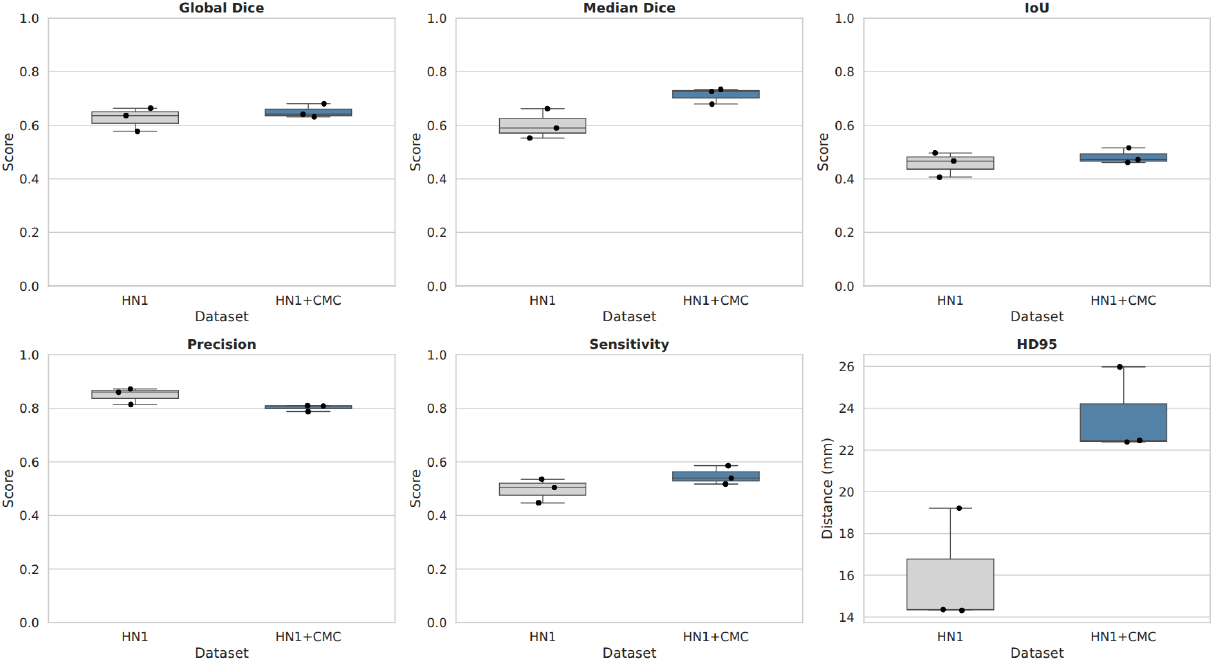
Boxplots summarizing segmentation performance for HN1 only vs HN1+CMC across three folds. The first row shows overlap metrics (Global Dice, Median Dice, and IoU), while the second row shows classification and distance metrics (Precision, Sensitivity, and HD95)

The overlap metrics indicate that both Global Dice and Median Dice achieved consistently high agreement with the ground truth across folds, with mean values of 0.6504 and 0.7125, respectively. IoU followed a similar trend with slightly lower absolute values (0.4823), reflecting its stricter penalization of mismatches. These trends confirm that the model maintained reliable contour adherence and robust volumetric segmentation quality across folds.

### 4.4 Qualitative Evaluation

Beyond the quantitative performance, Figure 6 provides representative qualitative results on head and neck CT slices. The predicted tumor masks (green) demonstrate strong spatial agreement with the expert annotations (yellow), reflecting the model’s ability to capture tumor morphology and boundary details. In most cases, the contours overlap closely, confirming the network’s robustness and its capacity to generalize to unseen data. Instances of minor under-segmentation are primarily observed in regions with diffuse or irregular tumor boundaries, which is consistent with the relatively lower sensitivity values reported in Table 3. These observations complement the quantitative findings, highlighting the trade-off between conservative predictions and complete tumor coverage.

**Figure 6:**
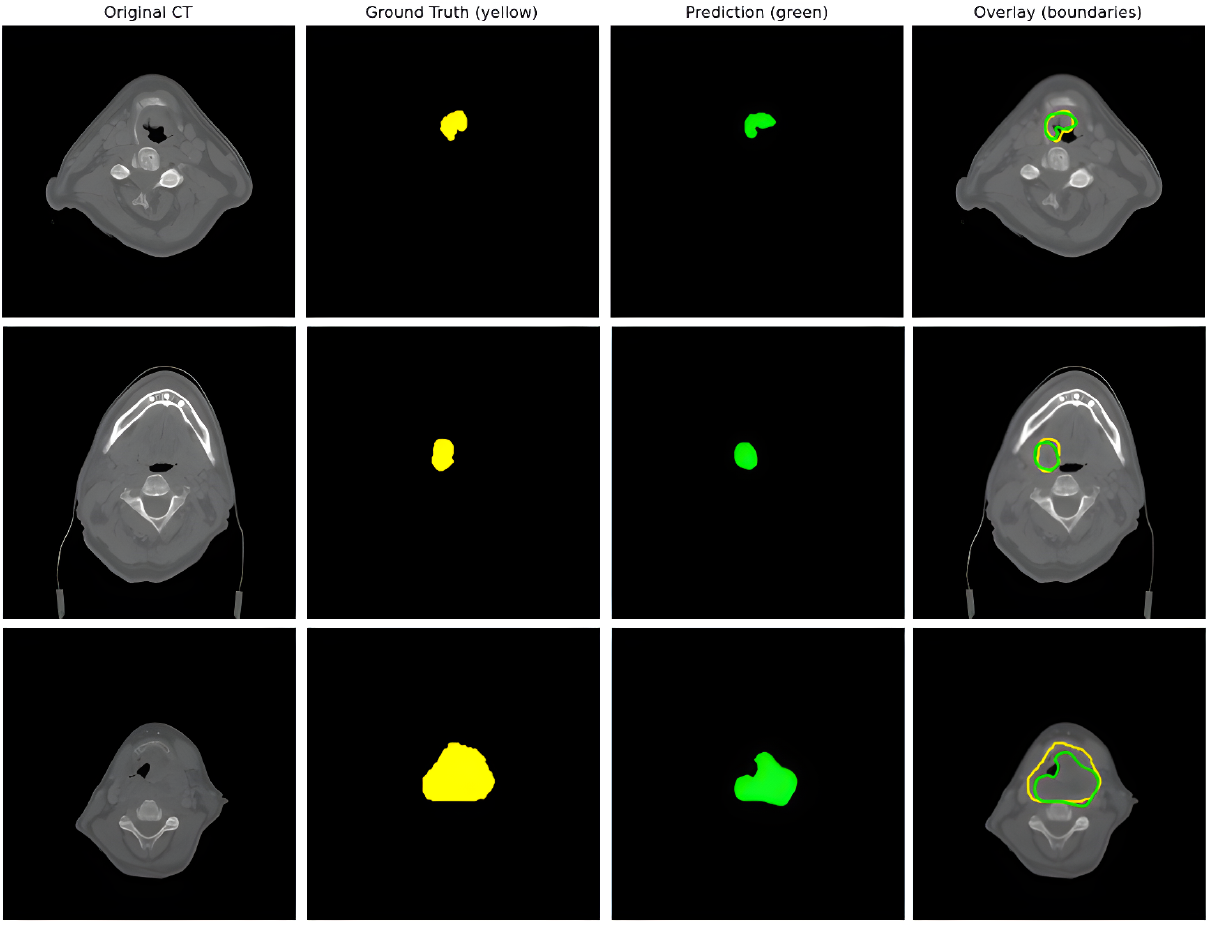
Qualitative visualization of segmentation results on a head and neck CT slice. Panels (a)–(d) show: original CT, ground truth mask (yellow), predicted mask (green), and overlay with solid boundaries (yellow = ground truth, green = prediction)

## 5 Discussion

### 5.1 Effect of adding private data

Integrating a modest number of private CMC cases improved overlap metrics on average, with the largest gains in Median Dice. Benefits were not uniform across folds, which suggests sensitivity to case mix and split composition. The experimental results indicate that incorporating 30 private CMC cases, which accounted for approximately 18% of the training data, influenced segmentation performance on the public HN1 dataset. Notably, Fold 1 demonstrated consistent improvements across several metrics, including Global Dice (0.6799 vs. 0.6633), Median Dice (0.6787 vs. 0.5894), IoU (0.5150 vs. 0.4962), and HD95 (22.38 mm vs. 14.32 mm). These findings suggest that limited additional training data from a complementary source can enhance volumetric overlap and boundary localization in certain folds.

### 5.2 Precision–Sensitivity Analysis

Across all folds, precision values (0.7876–0.8102) remained consistently higher than sensitivity values (0.5172– 0.5857), indicating a conservative segmentation strategy. This tendency minimized false positives, as evidenced by consistently high specificity (> 0.9998), but also led to underestimation of tumor regions, particularly in small or diffuse lesions. Such a trade-off is common in head and neck tumor segmentation tasks, where lesion boundaries are often irregular and challenging to capture in their entirety.

### 5.3 Boundary Accuracy

Boundary accuracy, assessed using HD95, demonstrated relatively stable performance across folds (22.38– 25.98 mm). The lowest HD95 value was achieved in Fold 1 (22.38 mm), which also corresponded to the highest Global Dice score. This concordance suggests that improved volumetric overlap was accompanied by more accurate boundary alignment, highlighting the benefit of leveraging complementary private data for refining contour precision.

Overall, the experiments demonstrate that augmenting public HN1 data with a modest number of private CMC cases can provide measurable performance improvements, particularly in terms of volumetric overlap and boundary accuracy. However, the benefits were fold-dependent and not consistently observed across all evaluation metrics, underscoring the influence of data distribution and limited private dataset size. These results emphasize the importance of integrating diverse datasets for head and neck cancer segmentation and suggest that larger, more balanced private cohorts may be required to achieve stable, generalizable gains across all folds and evaluation metrics.

## 6 Conclusion

This study demonstrates the potential of a 3D model for automated segmentation of head and neck cancer lesions from CT images. Unlike most existing approaches that rely on PET/CT, our work shows that CT-only models can achieve comparable performance, offering a more accessible and patient-friendly alternative given the lower cost and wider availability of CT imaging. Furthermore, while the majority of prior head and neck cancer segmentation studies employ 2D networks, our 3D framework captures richer spatial context and achieves robust results. Adding a private CMC cohort improved the overlap and robustness, but boundary accuracy remains challenging, and fold-to-fold variability suggests sensitivity to data composition. These observations indicate that even limited, complementary data can help generalization while highlighting center-specific differences that need attention.

Looking ahead, future work may focus on integrating domain adaptation techniques to reduce center-specific variability, leveraging few-shot or semi-supervised learning to mitigate limited annotation challenges, and systematically benchmarking CT-only models against PET/CT fusion approaches. Expanding private datasets to achieve a more balanced representation and reduce fold-dependent variability, incorporating advanced foundation models such as MedSAM or hybrid approaches that combine nnU-Net with bounding box-based localization, and exploring data augmentation strategies or multimodal imaging (e.g., PET/CT) could further enhance sensitivity, robustness, and clinical applicability across diverse patient populations.

## Data Availability

Public data used in this study available at https://www.cancerimagingarchive.net/collection/head-neck-radiomics-hn1/

## References

[1] Charlotte L. Brouwer, Roel J.H.M. Steenbakkers, and et al. Observer variation in delineation of target volumes in head and neck cancer. Radiotherapy and Oncology, 117(3):452–457, 2015.

[2] Mu Feng, Gilmer Valdes, and et al. Deep learning for head and neck cancer radiation therapy: A review. International Journal of Radiation Oncology*Biology*Physics, 107(4):766–778, 2020.

[3] Hyuna Sung, Jacques Ferlay, Rebecca L. Siegel, and Laversanne et al. Global cancer statistics 2020: Globocan estimates of incidence and mortality worldwide for 36 cancers in 185 countries. CA: A Cancer Journal for Clinicians, 71(3):209–249, 2021.

[4] Florian Dammann, Friedrich Bootz, Mathias Cohnen, Stefan Haßfeld, Marcos Tatagiba, and Sabrina Kösling. Diagnostic imaging modalities in head and neck disease. Deutsches Ärzteblatt International, 111(23-24):417–423, 2014.

[5] Ke Men, X Chen, Y Zhang, and et al. Deep deconvolutional neural network for target segmentation of nasopharyngeal cancer in planning ct images. Frontiers in Oncology, 7:315, 2017.

[6] Bulat Ibragimov and Lei Xing. Segmentation of organs-at-risks in head and neck ct images using convolutional neural networks. Medical Physics, 44(2):547–557, 2017.

[7] K He, X Zhang, S Ren, and J Sun. A deep learning model to automate skeletal muscle area measurement on ct images. Clinical Cancer Research, 25(12):3684–3692, 2019.

[8] Fabian Isensee, Paul F. Jaeger, Simon A.A. Kohl, Jens Petersen, and Klaus H. Maier-Hein. nnu-net: a self-adapting framework for u-net-based medical image segmentation. Nature Methods, 18:203–211, 2021.

[9] Valentin Oreiller, Vincent Andrearczyk, and Jreige et al. Head and neck tumor segmentation in pet/ct: The hecktor challenge. Medical Image Analysis, 77:102336, 2022.

[10] Benjamin Berthon, Michael Evans, Chris Marshall, Navin Palaniappan, Nick Cole, Vijay Jayaprakasam, Thomas Rackley, and Emiliano Spezi. Head and neck target delineation using a novel pet automatic segmentation algorithm. Radiotherapy and Oncology, 122(2):242–247, 2017.

[11] Yngve Mardal Moe and Groendahl et al. Deep learning-based auto-delineation of gross tumour volumes and involved nodes in pet/ct images of head and neck cancer patients. European Journal of Nuclear Medicine and Molecular Imaging, 48:2782–2792, 2021.

[12] Zhi Guo, Ning Guo, Ke Gong, Quanzheng Li, et al. Gross tumor volume segmentation for head and neck cancer radiotherapy using deep dense multi-modality network. Physics in Medicine & Biology, 64(20):205015, 2019.

[13] Jintao Ren, Jesper Grau Eriksen, Jasper Nijkamp, and Stine Sofie Korreman. Comparing different ct, pet and mri multi-modality image combinations for deep learning-based head and neck tumor segmentation. Acta Oncologica, 60(11):1399–1406, 2021.

[14] Mohammed A. Naser and Khurram A. Wahid. Head and neck cancer primary tumor auto segmentation using model ensembling of deep learning in pet/ct images. In 3D Head and Neck Tumor Segmentation in PET/CT Challenge, Lecture Notes in Computer Science, pages 121–133. Springer, 2021.

[15] Skylar S. Gay, Carlos E. Cardenas, Callistus Nguyen, Tucker J. Netherton, Cenji Yu, Yao Zhao, Stephen Skett, et al. Fully automated, ct-only gtv contouring for palliative head and neck radiotherapy. Scientific Reports, 13(1):21797, 2023.

[16] Juha Liedes, Hannu Hellström, Otto Rainio, et al. Automatic segmentation of head and neck cancer from pet-mri data using deep learning. Journal of Medical and Biological Engineering, 43:532–540, 2023.

[17] Lin M. Zhao, H. Zhang, D. D. Kim, et al. Head and neck tumor segmentation convolutional neural network robust to missing pet/ct modalities using channel dropout. Physics in Medicine and Biology, 68(9):095011, 2023.

[18] F. C. J. Reinders, M. H. F. Savenije, M. de Ridder, et al. Automatic segmentation for magnetic resonance imaging guided individual elective lymph node irradiation in head and neck cancer patients. Physics and Imaging in Radiation Oncology, 32:100655, 2024.

[19] Loo Wee and Andre Dekker. Data from head-neck-radiomics-hn1. The Cancer Imaging Archive, 2019. Available at 10.7937/tcia.2019.8kap372n.

[20] Fabian Isensee, Paul F. Jaeger, Simon A. A. Kohl, Jens Petersen, and Klaus H. Maier-Hein. nnu-net: a self-configuring method for deep learning-based biomedical image segmentation. Nature Methods, 18(2):203–211, 2021.

